# The COSEVAST Study: Unravelling the role of Arterial Stiffness in COVID-19 Disease severity

**DOI:** 10.1101/2020.12.18.20248317

**Authors:** Sanjeev Kumar, Neeraj Kumar, Abhyuday Kumar, Divendu Bhushan, Amarjeet Kumar, Ajeet Kumar, Veena Singh, Prabhat Kumar Singh

## Abstract

**Background:** Based on the detailed review of available research and case studies reported in reputed international journals, it can be concluded that endothelial damage (Endotheliitis) both in small and large arteries may be an important factor of morbidity and mortality in COVID-19 patients. Arterial stiffness due to Endothelial Dysfunction has been established as an independent and specific marker of various chronic cardiovascular diseases.

**Objective:** Our objective was to examine functional impairment of the arteries in COVID-19 disease and establish the non-invasive measurement of Arterial Stiffness as an independent marker of disease severity.

**Methods:** We recorded the Arterial Stiffness of 23 Mild, 21 Moderate and 20 Severe COVID-19 patients grouped on latest NIH severity criteria. We observed Arterial Stiffness of COVID-19 patients with standard parameters like non-invasive Carotid-Femoral Pulse Wave velocity (cfPWV), Age-Normalized increase in cfPWV (ANI_cfPWV).

**Results:** Moderate and Severe COVID-19 patients have extremely elevated arterial stiffness than Mild patients. In Mild patients, cfPWV (829.1 ± 139.2 cm/s) was extremely significantly lower than both Moderate (1067 ± 152.5 cm/s, P< 0.0001) and Severe (1416 ± 253.9 cm/s, P < 0.0001) patients. ANI_cfPWV in Moderate and Severe patients was significantly higher than Mild patients. (Mild: 101.2 ± 126.1 cm/s; Moderate: 279 ± 114.4 cm/s; Severe: 580.1 ± 216.4 cm/s; intergroup P <0.0001). Conclusion: Our findings strongly suggest that arterial stiffness can be an independent and accurate marker for objective risk stratification and therapeutic alleviation of the acute cardiovascular complications like MODS in COVID-19.

## Background

The acute respiratory viral infection caused by severe acute respiratory syndrome coronavirus 2 (SARS-CoV-2) is named as Corona Virus Disease 2019 (COVID-19) and is declared as a pandemic by the World Health Organization (WHO). As of first week of December 2020, there are around 9.5 million confirmed cases of COVID-19 in India alone, including more than 130,000 deaths. There is a rise in the number of cases every day, affecting millions of people around the globe and resulting in a public health emergency. Moreover, medical professionals are finding it very difficult to manage the disease in terms of treatment options. Many drugs have been tried under emergency use authorization like Remdesivir or convalescent plasma without much success. Preventive measures like vaccines are being discovered and some are in advanced stages of testing.[1] There is an urgent need to stratify the risk, guide the therapy in a proper manner and control the short and long term complications like target organ damage. Although this disease primarily looks like an acute pulmonary disease, the manifestation of cardiovascular damage strongly suggests that there is an arterial involvement. Various studies have pointed to severe and acute endothelial damage in moderate to severe cases of COVID-19. There is a need to clearly establish relationship between the arterial damage and severity in such patients to improve understanding of the pathophysiology of the disease. Such evidence may radically change the therapeutic strategy or give rise to a new therapy to improve the prognosis and lessen the burden on the healthcare facilities to a large extent.

## Introduction

It has been extensively highlighted that SARS-CoV-2 affects the RAS (Renin-Angiotensin System) and the Angiotensin converting enzyme-2 receptors play most important role in presentation of the COVID-19 disease. [2-5] SARS-CoV-2 infects the host using the Angiotensin Converting Enzyme 2 (ACE2) receptor, which is expressed in several organs, including the lung, heart, kidney, brain and intestine. This is due to the fact that ACE2 is a type I membrane protein expressed on endothelial cells in the kidney, heart, gastrointestinal tract, blood vessels, and, importantly, lung AT2 alveolar epithelial cells, which are particularly prone to viral infection. In COVID-19, Sepsis is observed as the most important cause of target organ damage leading to morbidity and subsequent mortality.

As SARS-CoV-2 is a new coronavirus, and its cardiovascular complications and the underlying pathology is still emerg-ing. However, it is accepted that the virus affects the total vasculature in human body and this infection becomes a highly accelerated process for the target organ damage. In the case of acute cardiological manifestation, it is termed as Acute COVID-19 Cardiovascular Syndrome (ACovCS) by the American Heart Association’s white paper. [6]

European Society of Cardiology (ESC) has published a detailed review paper [7] on the involvement of various cardiovascular target organs in COVID-19 disease. This review clearly establishes a close two-way relationship between COVID-19 disease and all cardiovascular diseases (CVD). Even though clinical symptoms of the disease with initial infection show respiratory distress and severe pneumonia, in the later stages the complete cardiovascular system gets affected with direct and indirect damage to other organs dependant on it. The prognosis is even worse in the patients with pre-existing cardiovascular system involvement. Cardiovascular involvement in COVID -19 is seen as key manifestations. In particular, the endothelial cells or pericytes inflammation could lead to severe microvascular and macrovascular dysfunction. Every stage of COVID-19 disease which leads to heart or similar target organ failure shows arterial involvement, basically starting from endothelial cell involvement. In COVID-19, Sepsis is observed as the most important cause of target organ morbidity and subsequent mortality. In Sepsis, the severity is directly associated with Endothelial dysfunction. It may be deduced that pathophysiology of Sepsis is mainly dependent on level of integrity and functioning endothelium. Hence an objective assessment of Endothelial function may serve as a more accurate prediction tool and a target for therapies aimed at ameliorating Sepsis occurring in later stages of COVID-19.

The best way to assess Endothelial Dysfunction is assessment of its clinical manifestation, i.e. increase in the Arterial Stiffness. There have been multiple in vitro and in vivo studies which have shown that the vascular endothelium is an important factor in setting the vascular tone and endothelial dysfunction leads to arterial stiffness.[8] Arterial stiffness and enhanced wave reflections are markers of cardiovascular disease and independent predictors of cardiovascular risk.[9–12] Stiffening of the large arteries and enhanced wave reflections lead to increased left ventricular (LV) afterload, disturbed coronary perfusion, and mechanical fatigue of the arterial wall.[13]

The relationship between systemic inflammation and arterial stiffness is well established in the literature.[14] The cause-and-effect relationship that acute systemic inflammation leads to deterioration of large-artery stiffness. Given the importance of aortic stiffness for cardiovascular function and risk, the therapeutic interventions with anti-inflammatory properties will be potentially very useful in reducing the COVID-19 morbidity and mortality.

The relationship between systemic inflammation and arterial stiffness is well established in the literature.[14] The cause-and-effect relationship that acute systemic inflammation leads to deterioration of large-artery stiffness. Given the importance of aortic stiffness for cardiovascular function andrisk,thetherapeuticinterventionswithanti-inflammatory properties will be potentially very useful in reducing the COVID-19morbidityandmortality.

Findings in a study [15] on induced inflammation have shown that that an acute inflammation caused a temporary increase in central blood pressure and arterial stiffness in terms of Pulse Wave Velocity. This implies the increased risk of cardiovascular events associated with acute systemic inflammation in the COVID-19.

COVID-19 disease related worldwide research and proposed mechanisms pointed to patho-physiological involvement of endothelial dysfunction and arterial wall compromise. However, there was no empirical evidence of the functional compromise of arterial walls.

Hence, a study was urgently needed to study increase in Arterial Stiffness in COVID-19 patients due to systemic inflammation so as to stratify the risk and mitigate further cardiovascular damage with guided therapeutic treatment based on severity of arterial stiffness.

This pressing need is justified by a comprehensive review article [16] published after the present study was envisaged. We hope that the findings from our study will fulfill the need to a large extent.

## Materials and Methods

The present prospective non-randomized observational study titled - “To study the relationship of COVID-19 SEVerity with Arterial STiffness : A prospective cross sectional study” (‘COSEVAST study’) was conducted in the COVID-19 ICU, Medical ICU and various wards of dedicated Covid hospital at AIIMS, Patna, Bihar, India. The study protocol, informed consents and other trial-related documents received the written approval of Institutional Ethics Committee (IEC No. AIIMS/Pat/IEC/2020/595) AIIMS Patna,Bihar,India. The study design was registered with the Clinical Trials Registry of India (CTRI No. CTRI/2020/10/028489).

All COVID-19 patients were subject to RT-PCR test and had a confirmed infection of SARS-Cov-2 virus. Participants, after understanding the study protocol and procedures, gave their written informed consent for the study. The exclusion criteria were known history of any of these diseases - Diabetes Mellitus (DM), Hypertension (HTN), CAD, Stroke, Neuropathy, PAD, Nephropathy, MI, Pregnancy, Peripheral Oedema or inflammation, Cardiac arrhythmia and any pre-existing Cardiovascular disorder.

### Patient Categorization

The selected patients after fulfilling the inclusion criteria were grouped into three categories – Mild, Moderate and Severe category based on latest NIH Guidelines [17] as-

A. Mild category: Individuals with mild signs and symptoms like fever, cough, sore throat, malaise, headache, muscle pain, nausea, vomiting, diarrhea, loss of taste and smell but who do not have shortness of breath, dyspnea, or abnormal chest imaging.

B. Moderate category: Individuals who show evidence of lower respiratory disease during clinical assessment or imaging and who have saturation of oxygen (SpO2) 94% on room air at sea level and

C. Severe category: Individuals who have saturation of oxygen (SpO2) <94% on room air at sea level, a respiratory rate of >30 breaths/min, PaO2/FiO2 <300 mmHg, or lung infiltrates >50%.

The range of treatment protocols followed in this study were based on our institutional standard operating procedure (SOP) for the management of COVID-19 patients based on their clinical severity category and it included antivirals (Remdesivir), broad spectrum antibiotics, low molecular weight heparin (Enoxaparin) immunomodulators (steroids, Tocilizumab) and supportive management (oxygen via nasal cannula, face mask, non-rebreathing face mask; non-invasive or invasive mechanical ventilation; awake proning).

### Measures of Arterial Stiffness

With the increased awareness about Arterial Stiffness during pre-Covid era, clinicians have started adding measurement and treatment of underlying arterial stiffness in their clinical practice.[18] International healthcare societies like the European Society of Hypertension (ESH) and European Society of Cardiology (ESC) have included PWV measurement in their 2003 guidelines for management of hypertension.[19,20]

Arterial stiffness assessed by PWV (pulse wave velocity) correlates to the number of treated and non-treated cardiovascular risk factors, atherosclerotic events and cardiovascular risk as predicted by the Framingham risk equations.[21] PWV is also positively correlated with carotid intima media thickness, a marker of atherosclerotic burden in the coronary arteries.[22] In this study, Arterial stiffness is primarily measured in terms of pulse wave velocity, which is a recognized gold standard.

Pulse wave velocity, which is a relevant indicator of arterial stiffness, can be measured non-invasively with a variety of devices. However, since COVID-19 is a highly contagious disease, a fully automatic device with minimal contact, lesser exposure time and proximity with the patient need to be maintained.[23] It would be always challenging for observer to conduct the entire test by wearing full Personal Protection Equipment (PPE) kit. So a test device which avoids close proximity, longer exposure and holding a probe while testing, was needed for the study. We found that the medical device PeriScope™(Manufactured by M/s. Genesis Medical Systems Pvt. Ltd., Hyderabad, India) was suitable as it fulfilled all the above requirements. PeriScope is a clinically validated and tested the non-invasive medical device used to measure Brachial Ankle PWV (baPWV) and derives the Carotid Femoral PWV (cfPWV), which is equivalent to aortic PWV.[24] A population-based study with 3969 subjects using ‘PeriScope’ has established the role of arterial stiffness in various cardiovascular diseases (CVD).[25] Aortic pressure values also have been established as surrogate markers for arterial stiffness. PeriScope estimates the Aortic Pressures and the Systolic Pressure Augmentation (AugP) at the root of the Aorta due to the increased arterial stiffness. The Pressure Augmentation Index (AIx) values found by PeriScope were compared with other internationally accepted non-invasive device and found to be very accurate and highly comparable.[26] All the tests were conducted as per the standard procedure given in the operator’s manual of PeriScope. Electrocardiogram (ECG) Electrodes were placed on ventral surface of both wrists and medial side of ankles, and BP cuffs were wrapped on both upper arm brachial artery and tibial artery above ankles. All the ECG and pressure recordings were done automatically and data were stored in the personal computer for analysis. The built in validated proprietary algorithm within the PeriScope PC software calculated the following parameters from the waveforms, which were stored in the computer for analysis like systolic blood pressure (SBP), diastolic blood pressure (DBP), mean arterial pressure (MAP), pulse pressure (PP), heart rate (HR), carotid-femoral pulse wave velocity (cfPWV).

### Data Analysis

Data are expressed as mean ± SD. Statistical analysis was performed using the GraphPad PRISM software version 5.03 (GraphPad software Inc., San Diego, California, USA). Difference between all the groups were evaluated by unpaired Student’s ‘t’ test and ANOVA. Linear regression analysis and Pearson’s correlation analysis were performed to evaluate the association between various confounding factors to rule out interdependence. Probability values of P < 0.05 were considered to indicate statistical significance. All P values are two-tailed.

## Results

Total 64 patients (42M/22F) patients were recruited in the study. The patient demographic variables were shown in Table 1. All the patients were under the healthy and normal BMI category. Since the patients did not have any known comorbidity, Lipid profile or Blood sugar levels were not taken into consideration. The minimum age was 18 years and maximum was 69 years. Although there was a difference between the mean values of Age, Weight and height; but it was statistically not significant between the groups. Figure 2 shows the graphical representation of overall patient demographics as age, weight, height and BMI as (Figure 2a, b, c and d).

**Fig. 1.**
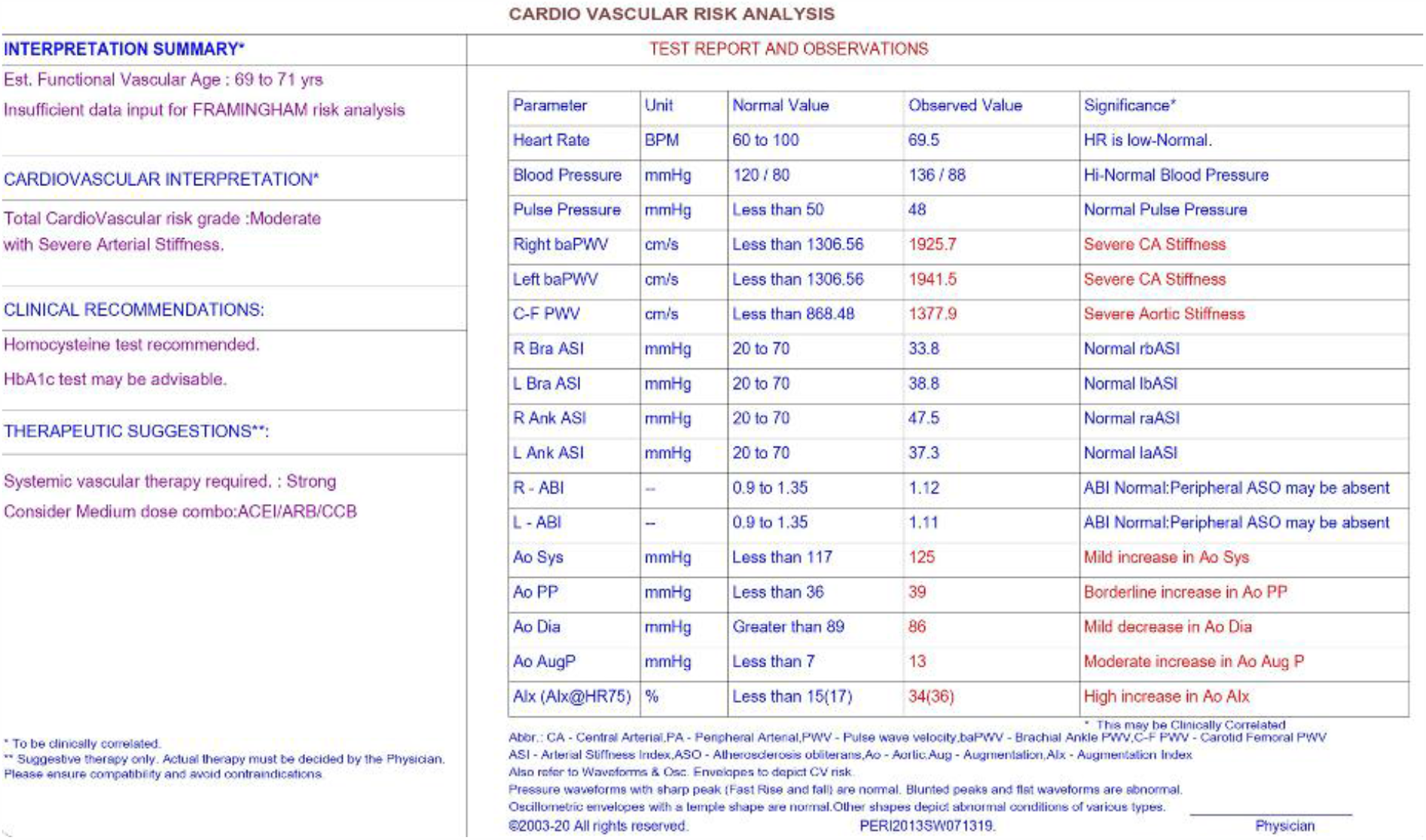
Typical PeriScope test report of a Severe Patient in the COVID-19 ICU

**Fig. 2:**
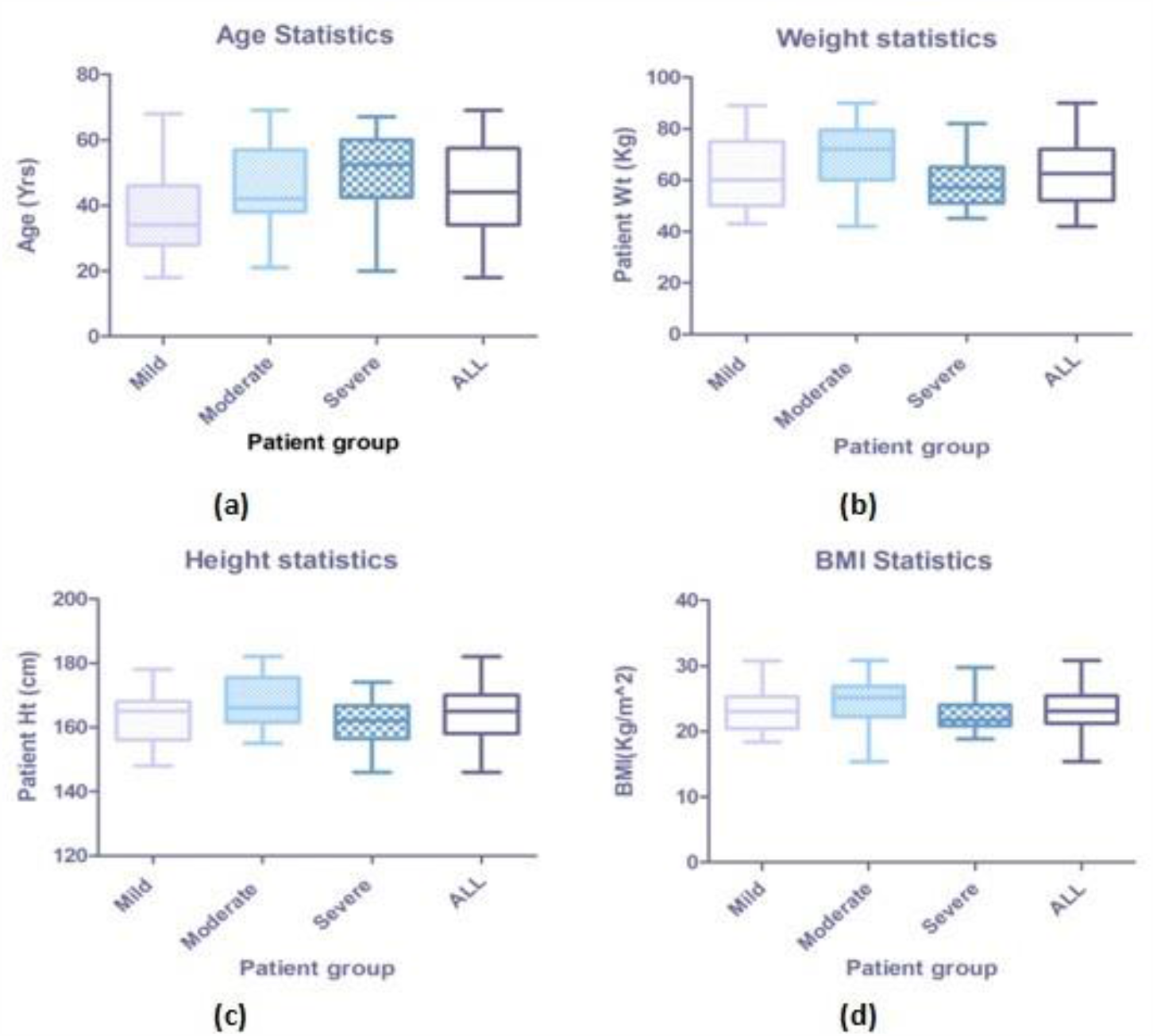
Patients Demography a. Age, b. Weight, c. Height, d. BMI

Although Arterial Blood Gas (ABG) Analysis was carried out for all the patients, it was not possible to find the point-of-time PaO2% value of each patient at the time of arterial stiffness test. For this reason, stat SpO2% reading was taken as the measure of Oxygen saturation for accurate comparison. Also, since it was not ethically correct[27] and clinically possible to check oxygen saturation without or by removing external Oxygen support in all cases, the best way to quantify the severity was taking ratio of SpO2% and FiO2% as an indication of respiratory distress. Earlier studies and guidelines suggest SpO2% to FiO2% (SF) ratio is a reliable noninvasive surrogate for PaO2% to FiO2% (PF) ratio [28-30] especially in COVID-19 patients. All of the Severe patients were admitted into the COVID-19 ICU whereas all of the Mild patients were admitted into COVID-19 and General wards. Some of the Moderate patients were in the ICU and some were in the General ward with Oxygen support. The statistical analysis of the segregation criteria is as shown in Table 2. The Patient categorization can be clearly visualized by graphical representation (Figure 3) of the data. The Figure 3a shows statistical analysis of respiration rate of the three groups, Figure 3b shows FiO2%, Figure 3c shows SpO2% whereas Figure 3d shows the statistical analysis of SF ratio (SpO2/FiO2) respectively.

**Table 1.** Demographic characteristics as per COVID-19 severity in the present study

| Sr. No. | Parameter | Mild(1) | Moderate(2) | Severe(3) | All | P-value |  |  |
| --- | --- | --- | --- | --- | --- | --- | --- | --- |
|  |  |  |  |  |  | Grp1-2 | Grp2-3 | Grp1-3 |
| 1 | Sex(M/F) | 13/10 | 18/3 | 11/9 | 42/22 | — | — | — |
| 2 | Age(yrs) | 37.87±13.36 | 45.29±13.07 | 50.50±12.56 | 44.25±13.85 | 0.0701 | 0.2007 | 0.0028* |
| 3 | Wt(Kg) | 61.96±13.32 | 70.14±12.75 | 59.05±10.12 | 63.73±12.89 | 0.0438* | 0.0038* | 0.4307 |
| 4 | Ht(cm) | 162.8±8.574 | 167.9±8.402 | 161.7±7.680 | 164.1±8.555 | 0.0541 | 0.0174* | 0.6403 |
| 5 | BMI(Kg/m2) | 23.18±3.547 | 24.78±3.492 | 22.47±2.532 | 23.48±3.334 | 0.1405 | 0.0207* | 0.4605 |

Values are expressed as Mean±SD. \*Significant difference in mean values.

**Table 2.** Categorization criteria as per COVID-19 severity in the present study

| Sr. No. | Parameter | Mild(1) | Moderate(2) | Severe(3) | P-value |  |  |
| --- | --- | --- | --- | --- | --- | --- | --- |
|  |  |  |  |  | Grp1-2 | Grp2-3 | Grp1-3 |
| 1 | Resp.rate(BrPM) | 20.87±1.180 | 22.33±4.139 | 29.55±13.19 | 0.1113 | 0.0219* | 0.0031* |
| 2 | FiO2(%) | 21.00±0.0 | 35.48±12.68 | 57.85±23.13 | <0.0001*\$ | 0.0004* | <0.0001*\$ |
| 3 | SpO2(%) | 97.30±1.428 | 96.48±2.136 | 96.50±2.236 | 0.1350 | 0.7994 | 0.0861 |
| 4 | S/Fratio | 463.4±6.801 | 304.1±101.6 | 195.9±82.33 | <0.0001* | 0.0006* | <0.0001* |

Values are expressed as Mean±SD. \*Significant difference in mean values. \$Mann-Whitney 't' test

**Fig. 3.**
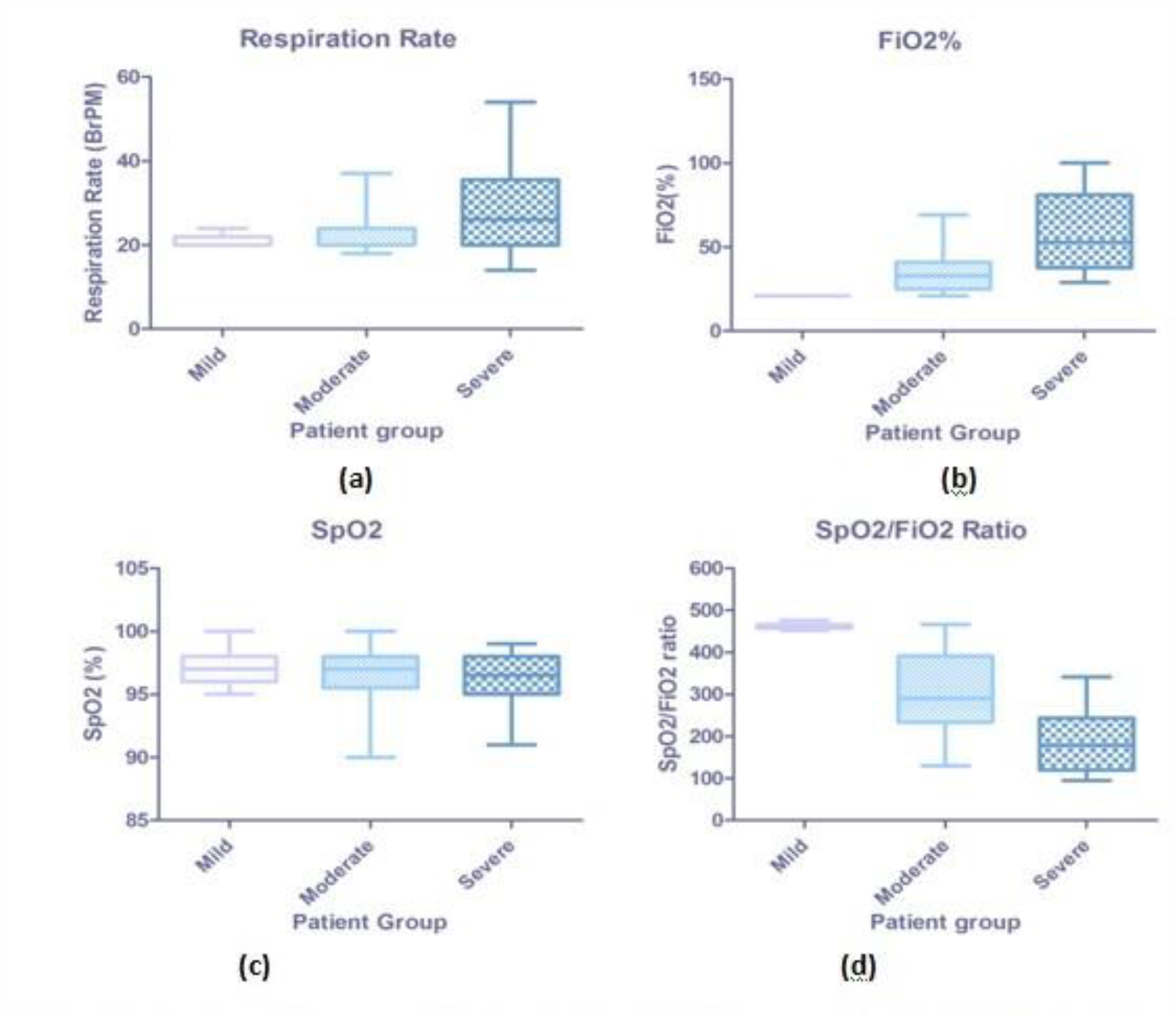
Patient Categorization a. Respiration Rate, b. FiO2%,c. SpO2%, d. SpO2%/FiO2% ratio

As the ‘normal’ value arterial stiffness is a function of a person’s chronological age [31], (i.e. the normal value of Arterial stiffness goes on increasing with age), the real measure of abnormal increase in Arterial stiffness is the difference of the ‘predicted’ cfPWV value of a ‘normal’ subject of that age and actually measured cfPWV. Hence, Arterial Stiffness was analyzed using the values of Carotid Femoral Pulse Wave velocity (cfPWV) and increase in cfPWV from the age dependent normal value. Also, in our study, the regression analysis between cfPWV and HR showed very low correlation (Multiple r = 0.2180 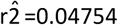, P=0.0835). Hence, the effect of HR on cfPWV values is negligible. It can very safely be stated that the findings about arterial stiffness from the study are independent of effect of HR. No other parameters like Heart rate or Pulse Pressure showed consistent increase in their mean values between Mild, Moderate and Severe groups. Heart Rate (Mild: 76.06 ± 14.47 BPM, 95% CI 69.81 to 82.32; Moderate:78.20± 17.20 BPM, 95% CI 70.36 to 86.03; Severe: 98.07± 18.58 BPM 95% CI 89.37 to 106.8), Brachial Pulse Pressure (Mild: 38.04 ± 7.957 mmHg, 95% CI 34.60 to 41.48; Moderate: 45.95 ± 9.113 mmHg, 95% CI 41.80 to 50.10; Severe: 42.40 ± 12.09 mmHg, 95% CI 36.74 to 48.06) respectively. Refer Fig. 4 for the graphical comparison.

**Fig. 4.**
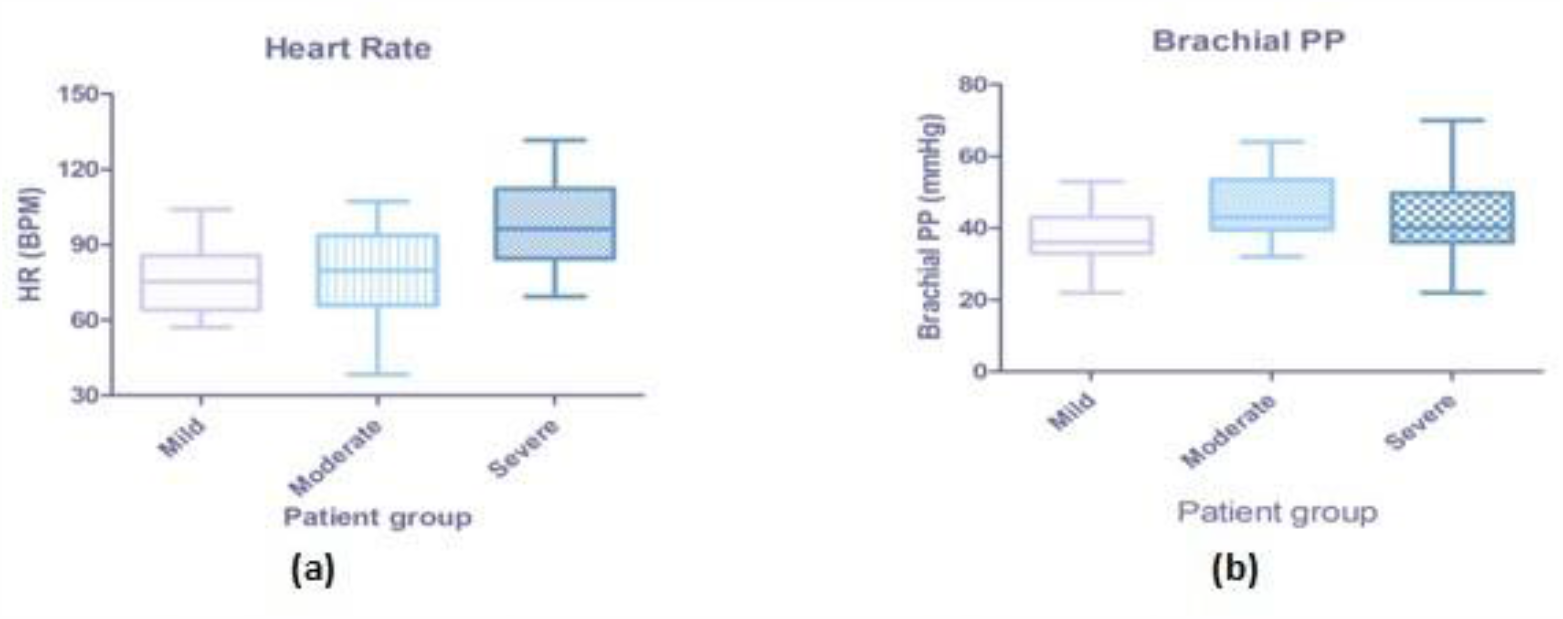
Related measurements in the study - a. Heart Rate, b. Brachial Pulse Pressure

On the other hand, the differences in the measurements of Arterial Stiffness parameters were extremely significant in all three groups. All the Arterial stiffness measurements showed uniformity in trends and almost same rise as per the severity of COVID-19 disease. More importantly, since no confounding factor or dependent parameter was found in the statistical analysis of the data, Arterial Stiffness measures stood out as independent and consistent indicators of severity grade. The analysis of Arterial Stiffness parameters is as given in Table 3. These results can be properly visualized in comparison with the earlier graphs. Figure 5 gives a clear idea about the extremely statistically significant elevation of all Arterial Stiffness measurements across patient groups. The Figure 5a shows values of cfPWV in all three patient categories. Figure 5b shows Age Normalized increase in cfPWV.

**Table 3.** Patient Arterial Stiffness measures as per COVID-19 severity in the present study

| Sr. No. | Parameter | Mild(1) | Moderate(2) | Severe(3) | P-value |  |  |
| --- | --- | --- | --- | --- | --- | --- | --- |
|  |  |  |  |  | Grp1-2 | Grp2-3 | Grp1-3 |
| 1 | cfPWV(cm/s) | 829.1±139.2 | 1067±152.5 | 1416±253.9 | <0.0001* | <0.0001* | <0.0001* |
| 2 | cfPWVincrease#(cm/s) | 101.2±126.1 | 279±114.4 | 580.1±216.4 | <0.0001* | <0.0001* | <0.0001* |

Values are expressed as Mean±SD. \*Significant difference in mean values. #Age Normalized.

**Fig. 5.**
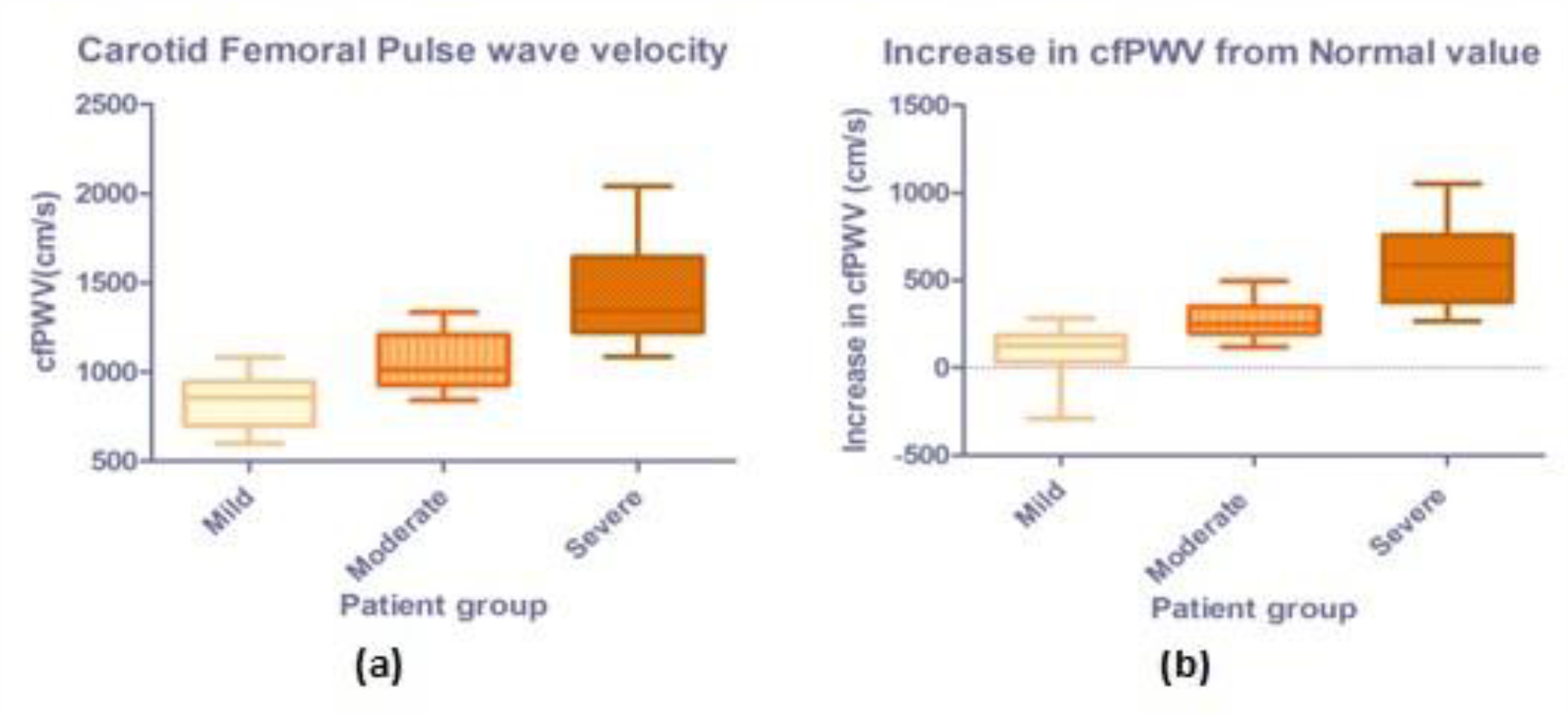
Arterial Stiffness measurements of COVID-19 Patients a. cfpwv, b. Age normalized increase in cfpw

## Discussion

The SARS-CoV-2 virus uses the same system of cell entry, which is triggered by binding of the viral spike (S) protein to angiotensin-converting enzyme 2 (ACE2) on the surface of the host cell. It is clearly evident that the full endothelial layer is affected throughout the body and the disease called Endotheliitis sets in due to COVID-19 infection. The Lancet article[33] shows clear images with endothelial layer involvement. Chen T et al [34] in their epidemiological studies show that COVID-19 patients admitted to hospital or to an intensive care unit (ICU) present frequently with accompanying conditions such as advanced age, hypertension, diabetes, and cardiovascular diseases implying compromised endothelial integrity may be playing a major part in COVID-19 mortality. Acute bilateral limb ischaemia (ALI) was observed in young and seemingly healthy patients without any history of atherosclerosis, atrial fibrillation, or preexisting blood clotting disorders indicating that COVID-19 infection might increase the incidence of ALI and be associated with poorer surgical results owing to the associated acquired hypercoagulability. [35] Explaining the potential role of Endothelial Dysfunction and endothelial layer compromise in COVID-19, the article in Cardiovascular Research [36] shows that Chronic Endothelial Dysfunction may lead to acute vascular inflammation under the attack of SARS-CoV-2 Virus. Prior chronic Endothelial impairment and/or direct viral infection of endothelial cells may work in tandem into a breakdown of endothelial function. Since endothelial layer is present everywhere throughout the vascular system in non-fenestrated form (in the arteries, lungs or the Heart), continuous non-fenestrated form (in the kidneys) and in the discontinuous form (in the liver), the acute endothelial function failure may lead to pathogenesis of sudden severe conditions in various organs. It may manifest as pneumonia and ARDS in the respiratory system, induce microcirculation disorders/myocardial injury in the heart, translate into activation of the coagulation pathway causing thrombosis or develop into tissue damage in the brain and in the Kidneys. The over-all vasculature is affected very quickly by this disease. The cfPWV which we observed in severe COVID-19 patients was higher than the cfPWV in patients with long term chronic diseases like ESRD, CAD, DM etc. For example, in CAD patients, it was 1204 ± 301.8 cm/s. [24-25] This indicates that COVID-19 damage to the vasculature within less than 15 days is comparable to the damage caused by CAD in number of years. Our COSEVAST study had very strict exclusion criteria. All patients with known Diabetes and Hypertension were excluded. Patients with all other cardiovascular diseases and related surgical intervention/ therapy were excluded. Patients with high obesity, history of smoking were also excluded. This was necessary to eliminate basal deviation of arterial Stiffness from the Normal vasculature. These exclusions reduced the total number of study subjects but removed any selection bias and amplified the validity of our findings. A similar large scale, longitudinal, multi-centric study (COVID-19 ARTErial Stiffness and vascular AgiNg CARTESIAN Study) [37] has been proposed and our study will try to follow harmonization with that study. Based on the findings of the present COSEVAST study, another interventional study will be undertaken in the near future to assess various drugs to reduce the abnormal increase of arterial stiffness in COVID-19 patients. The proposed study will be very helpful in establishing the most efficacious therapy in reducing COVID-19 complications. To the best of our knowledge, this is the first study to independently observe significant functional changes in vasculature due to arterial inflammation in COVID-19 patients in terms of non-invasive measures of Arterial stiffness.

## Limitations

As for the limitations, any of the patients did not abstain from the standard therapy to alleviate the COVID-19 disease symptoms since it was only an observational study. The sample size were very small. However, the therapy was uniform intra-group and did not include any drug which could alter the arterial stiffness of the patents. Especially, the therapy of Moderate and Severe groups was identical except the difference in Oxygen supplementation volume. We wanted to conduct multiple follow-up tests to find any trends in Arterial Stiffness, however it proved almost impossible to study patients with COVID-19 disease because of various reasons like changing status of the patient condition, restrictions on research staff movements and sometimes unfortunate death of the patients during the study. We are also in the process of undertaking a medium-term follow-up study of the patients in the present study after their discharge. These subjects will be observed for changes in Arterial Stiffness and occurrence of Cardiovascular disease, if any, over a period of 12 months with the PeriScope device.

## Conclusions

The results of this study establish non-invasive measurement of Arterial Stiffness as an independent severity marker in COVID-19 patients. It also strongly indicates that regular assessment and regulation of Arterial Stiffness by titration of therapeutics may reduce acute cardiovascular complications and mortality in COVID-19 patients significantly.

## Data Availability

NA

## Authors’ Contributions

Dr. Sanjeev Kumar: Supervision, Project administration, writing– review editing

Dr. Neeraj Kumar: Conceptualization, data curation, formal analysis, writing – original draft, writing– review editing Dr. Abhyuday Kumar: Methodology, data curation, writing– review editing

Dr. Divendu Bhushan: Methodology, data curation, writing– review editing

Dr. Amarjeet Kumar: Methodology, data curation, writing– review editing

Dr. Ajeet Kumar: formal analysis, writing– review editing Dr. Veena Singh: writing– review editing

Dr. Prabhat K Singh: Conceptualization supervision, visualization.

## Research in context

### A. Evidence before this study

The exact nature of the SARS-CoV-2 viral infection and its manifestation as COVID-19 disease is a matter of an ongoing discussion. Based on prior knowledge, it can be safely stated that COVID-19 is not solely a Respiratory system disease involving lungs. It also affects other bodily systems with target organ damage like Cardiovascular (Heart), Nervous system (Brain), Digestive (Bowels) as well as Renal (Kidneys). The common factor in all these systems is the vascular Endothelial layer in various forms. Pathological and clinical case study findings have shown Endotheliitis pervasive in COVID-19 cases. This leads to the hypothesis that acute functional compromise of vasculature leads to failure of various systems and organs in severe COVID-19 patients. Control and reversal of this damage may reduce the morbidity and mortality risk.

### B. Added value of this study

Although there is some pathological evidence of structural damage to the endothelial layer in COVID-19 patients, the systemic functional deterioration in the vasculature has not been studied. Our study is first of its kind to prove the functional deterioration of Arterial system in terms of abnormal increase in arterial stiffness as the probable cause of severity in COVID-19. Our findings strongly suggest that arterial stiffness can be an independent and accurate marker for objective risk stratification and therapeutic alleviation of acute cardiovascular complications like MODS (multiple organ distress syndrome) in COVID-19.

### C. Implications of all the available evidence

The basis of our study and its findings suggest that arterial stiffness measurement can be added as a simple and accurate measurement for objective risk stratification in COVID-19 patients. Early detection, monitoring and therapeutic control of Arterial Stiffness in COVID-19 patients may reduce acute cardiovascular complications. Arterial stiffness may be used as an independent marker for treatment and prognosis in COVID-19 patients.

